# Integrated Bile Acid Profile Analysis for The Non-invasive Staging Diagnosis of Ulcerative Colitis

**DOI:** 10.1101/2022.03.14.22272391

**Authors:** Lijuan Cao, Yun Wang, Lu Zhang, Yuan Che, Hai Huang, Hong Shen, Haiping Hao

## Abstract

Clinical staging diagnosis and progression tracking for ulcerative colitis (UC) is challenging as poor patient compliance of endoscopic biopsy, we aimed to explore a non-invasive integrative biochemical index to quantitative track and monitor pathological activity. Here we perform a study that integrates bile acid metabolomic profiling, metagenomic sequencing and clinical monitoring on serum and feacal samples from 24 active-state UC patients, 25 remission-state UC patients and 20 healthy volunteers from China. Besides known associations of *Fusobacterium nucleatum* and *Peptostreptococcus stomatis* with UC, we found several bile acid-transforming species, including 7α-dehydroxygenase and 7α/β-dehydrogenase expressing microbiota, were significant correlated with UC pathological activity. We identified 7 microbial gene markers that differentiated active and remission-stage UC and healthy control microbiomes. Relevantly, decreased serum deoxycholic acid /cholic acid ratio and increased fecal ursodeoxycholic acid/chenodeoxycholic acid ratio were associated with pathological activity of UC. Moreover, receiver operating characteristic analysis based on serum/fecal bile acids ratios was much accurate in prediction of active and remission stage outcome. This species-specific temporal change and bile acid dysregulation pattern linked to disease severity indicating that integrated microbiome-bile acid profile maybe implied for disease activity prediction, and that targeting microbiome-mediated restoring gut flora and bile acids homeostasis may be implicative of therapy efficacy. Collectively, these insights will help improve clinical diagnosis and optimize existing medical treatments.

## INTRODUCTION

Ulcerative colitis (UC) is a kind of chronic inflammatory bowel disease (IBD). Clinical practice of staging diagnosis and prognosis for ulcerative colitis (UC) is challenging as poor patient compliance with colonoscopy^1^. Biochemical test and biomarkers determination are fast and non-invasive ways of diagnosis, for instance, C-reactive protein (CRP) is one of the typical biochemical indictors of UC^2^, and merging auxiliary diagnostic biomarkers for the clinical monitoring of UC also includes erythrocyte sedimentation rate (ESR), feacal calprotectin (FC), stool lactoferrin and systemic microRNA expression profiling^2-4^. However, current clinical biochemical indexes can hardly applied for diagnosis and staging independent from endoscopy^5^, and the gold standard for non-invasive UC diagnosis is undefined^5^.

The pathogenesis of UC is complex and is involved in various factors, such as barrier function impairment, immune response in the lamina propria and luminal factors including intestinal microbiota^5^. Though advances have been made in comprehending the pathophysiology of UC, the exact etiology is still vague. Decreased colonic mucin 2 (MUC2) synthesis and impaired barrier function were observed in UC patients^6,7^. Genetic defect and dietary factors contribute to barrier impairment^8^. In addition, pro-inflammatory immune cells migrate from the lamina propria to the colonic mucosa and secret IL-13, TNF and IL-23, which are major proinflammatory cytokines involved in the UC progress^9,10^.

The dysbiosis of intestinal microbiota is closely associated with UC development. Reconstitution of intestinal microbial composition by fecal microbiota transplantation and probiotics administration leads to clinical remission^11^. *Akkermansia muciniphila*, which plays an important role in maintaining the intestinal barrier and reducing gut permeability, decreased in UC patients^12^. In addition, UC patients showed lower abundance of bacteria generating SCFAs, such as *Faecalibacterium prausnitzii* and *Roseburia hominis*^13^. Short chain fatty acids (SCFAs) are microbiota metabolites from polysaccharides degradation and exert the roles of relieving inflammation, protecting the intestinal barrier and enhancing antimicrobial host defense. For instance, butyrate drives the antimicrobial program in macrophages via inhibition of histone deacetylase 3 (HDAC3)^14^. Butyrate also suppresses macrophage inflammation through HDAC3 inhibition or G protein-coupled receptor 109A (GPR109A) activation^15,16^. Butyrate also decreases intestinal permeability and strengthens the intestinal barrier via an IL-10 receptor α subunit (IL-10RA)-dependent pathway^17^. Additionally, microbiota mediates the formation of the indole derivatives from dietary tryptophan. Indole-3-acetic acid (IAA) and indole-3-propionic acid (IPA) were reported to decrease in UC patients^18,19^. The indole derivatives are aryl hydrocarbon receptor (AhR) ligands and exert anti-inflammatory roles via the AhR−IL-22 signal and IL-10 signal^18-20^. *Lactobacillus reuteri* produces indole derivatives, which in turn activate AhR and reprogram intraepithelial CD4+ T cells into immunoregulatory T cells^21^. The microbiota-produced secondary bile acids which are deficient in ileal pouches of the patients undergoing colectomy, ameliorate inflammation partly dependent on G protein-coupled bile acid receptor 1 (TGR5)^22^. Although the dysbiosis of intestinal microbiota and the alternations of microbial metabolites influence the progression of UC, the relationship among single bacterial species, certain metabolites and the pathological mechanisms of UC is not fully clarified.

## RESULT

### Ulcerative colitis is associated with dramatic alternations in intestinal microbiota community structure

The alpha diversity showed that microbiota richness in UC active patients was significantly lower than healthy volunteers due to lower Shannon, Chao and Sobs index at the OTUs level (Figure 1A-D). Microbiota richness in UC remission patients was relatively higher than UC active patients but still lower than healthy volunteers (**Figure 1A-D**). Unweighed UniFrac distance-based principal coordinate (PCoA) analysis and non-metric multidimensional scaling (NMDS) analysis showed distinct clustering of intestinal microbiota communities for healthy volunteers, UC active patients and UC remission patients (**Figure 1E, Supplementary Figure S1A**). Distinct differences were observed in microbiota community structure between healthy volunteers and UC active patients (**Figure 1E, Supplementary Figure S1A**). The microbiota community structure of UC remission patients showed no obvious changes compared with either healthy patients or UC active patients (**Figure 1E, Supplementary Figure S1A**). The OTUs from Firmicutes and Bacteroidetes were the most enriched OTUs with a ternary plot visualization. In addition, proteobacteria OTUs were higher in UC active patients compared with healthy volunteer and Bacteroidetes OTUs tended to be higher in healthy volunteers (**Figure 1F**). The bar plot showed an overall view of the relative abundance of microbiota of all groups at phylum, class and order level. The changes at the phylum level showed abundance of Proteobacteria and Actinobacteria in UC patients was higher than healthy volunteers, whereas the abundance of Bacteroides was lower (**Figure 1G**). At the class level, the abundance of *Bacilli, Gamaproteobacteria* and *Actinobacteria* in UC patients was higher than healthy volunteers, whereas the abundance of *Clostridia, Bacteroidia* and *Negativicutes* was clearly lower (**Figure 1H**). At the order level, the abundance of *Lactobacillales, Enterobacteriales* and *Bifidobacteriales* in UC patients was higher than healthy volunteers, whereas the abundance of *Clostridiales, Bacteroidales* and *Selenomonadales* was clearly lower (**Figure 1I**). Additionally, the abundance of all the above bacteria in UC remission patients at phylum, class and order levels was between the abundance of healthy volunteers and UC active patients (**Figure 1G-J**). Differential analysis and LEfSe analysis exhibited *Bifidobacterium, Streptococcus* and *Enterococcus* in UC active patients took advantages while *Prevotella_9, Faecalibacterium, Eubacterium_rectale_group* and *Subdoligranulum* in healthy volunteers preponderant (**Figure 1K, Supplementary Figure S1B**). *Escherichia-Shigella* in UC active patients were higher than those in healthy volunteers (**Figure 1K, Supplementary Figure S1B**). In addition, differential analysis and LEfSe analysis showed *Streptococcus* and *Enterococcus* in UC active patients took advantages while *Faecalibacterium, Blautia, Prevotella_9, Eubacterium_rectale_group* and *Subdoligranulum* in UC remission patients were dominant (**Figure 1K, Supplementary Figure S1B**). UC patients, especially UC active patients, exhibit higher *Bifidobacterium, Streptococcus* and *Lactobacillales* levels than healthy volunteers (**Figure 1K, Supplementary Figure S1B**). This might result from the probiotics usage among UC patients, especially UC active patients who needed larger dose of probiotics treatment. Moreover, opportunistic pathogens, such as *Enterococcus* and *Escherichia-Shigella*, are obviously higher in UC active patients. In contrast, the abundance of these opportunistic pathogens is relatively lower in UC remission patients than in UC active patients, implying the improvement of the intestinal microbiota structure (**Figure 1L, Supplementary Figure S1C**). Moreover, compared with healthy volunteers, UC active patients has lower amount of *Faecalibacterium* and *Eubacterium_rectale_group*, which can produce short chain fatty acids (SFCAs) and benefit the intestinal microenvironment (**Figure 1N-Q**). Taken together, these results indicated active UC increases opportunistic pathogens abundance and reduced beneficial commensal bacteria and the dysbiosis partly recovers to normal along with UC remission.

**Figure 1.**
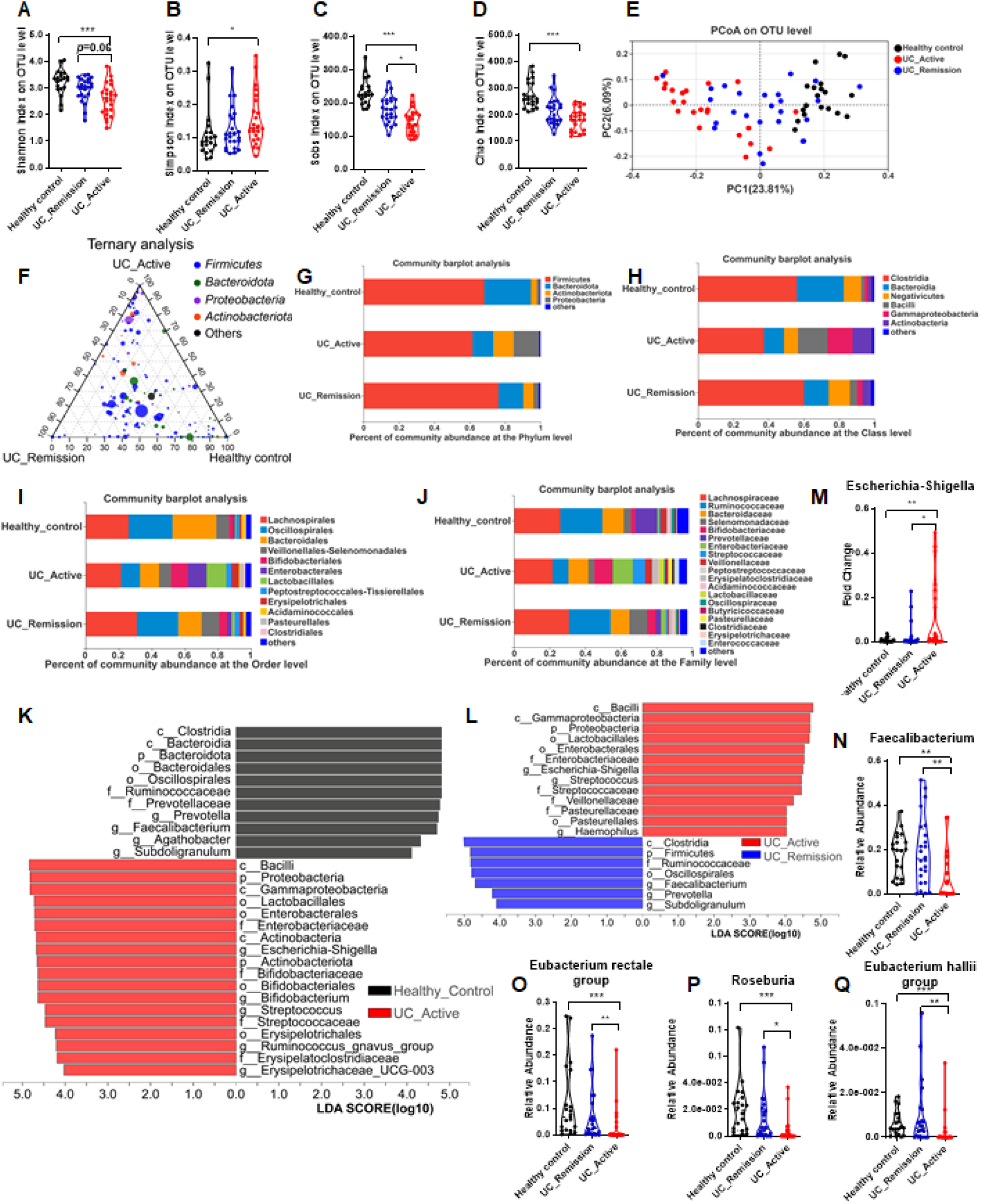
Ulcerative colitis is associated with dramatic alternations in intestinal microbiota community structure. (A-D) Alpha diversity in healthy controls, UC active and UC remission patients presented as Shanno (A), Chao1 (B), Sobs (C) and Simpson (B) index at OUT levels. (E) PCoA plot of unweighted UniFrac distances of microbiota composition in all groups at OTU levels. (F) Ternary plot of phylum-level OTUs from the most four abundant phylum in healthy controls, UC active and UC remission patients. (G-I) Relative abundance of microbiota of all groups at phylum (G), class (H) and order (I) level. Data are presented as bar plot with mean. (J) Significant taxa from phylum to genus levels identified by linear discriminant analysis (LDA) using LEfSe. Scores of the most differentially abundant taxons compared by healthy control and UC active. (K) Significant taxa from phylum to genus levels identified by LDA using LEfSe. Scores of the most differentially (H) abundant taxons compared by UC remission and UC active. (L-P) The relative abundance E. coli, Faecalibacterium, Eubacterium rectale group, Roseburia and Eubacterium hallil group in healthy control, UC active and UC remission patients analyzed by 16S rRNA sequencing. Data are presented as violin plots. ***p < 0.001, **p < 0.01, *p < 0.05; Kruskal-Wallis test.

### Bile acid disorder pattern is associated with pathological activity of UC

We previous disclosed bile acid regulation pattern in experimental UC model, and proved that bile acid dysregulation was interacted with pathological process of UC, herein, we intended to probe temporal change of metabolic regularity of bile acid in the pathology of UC in clinic. We analyzed a panel of major bile acids in serum and feces of 24 active-state UC patients, 25 remission-state UC patients and 20 healthy volunteers, we found that DCA/CA ratio were negatively and UDCA/CDCA were positively correlated with disease activity, in particular, DCA/CA were significantly distinctive between remission state and active state patients (**Figure 2A-K**). Previous studies have proposed that CA/CDCA ratio were positively related with inflammatory bowel diseases, here we also calculated serum and fecal CA/CDCA ratios, and found that, serum CA/CDCA ratio hardly changed in the progress of UC, while fecal ratio of CA/CDCA were quite positive in correlation with disease state (**Figure 2L and M**). We thus speculated that, gut microbiota mediated bile acid transformation is a hinge marker of UC, we further investigated other bile acid species which produced by microbiota, including total secondary/primary bile acids ratio and LCA/CDCA pathway, whereas, no significant correlation were found between these ratios and disease state (**Figure 2N-S**). Despite that total bile acid concentrations in serum and feces were hardly shift with disease progress of UC patients, and serum CRP were not responsive to remission or activity change (**Figure 2T-V**), our data indicated that specific metabolic pathway of bile acid, CA/DCA and CDCA/UDCA transformation, were interacted with disease process of UC (**Figure S2**).

**Figure 2.**
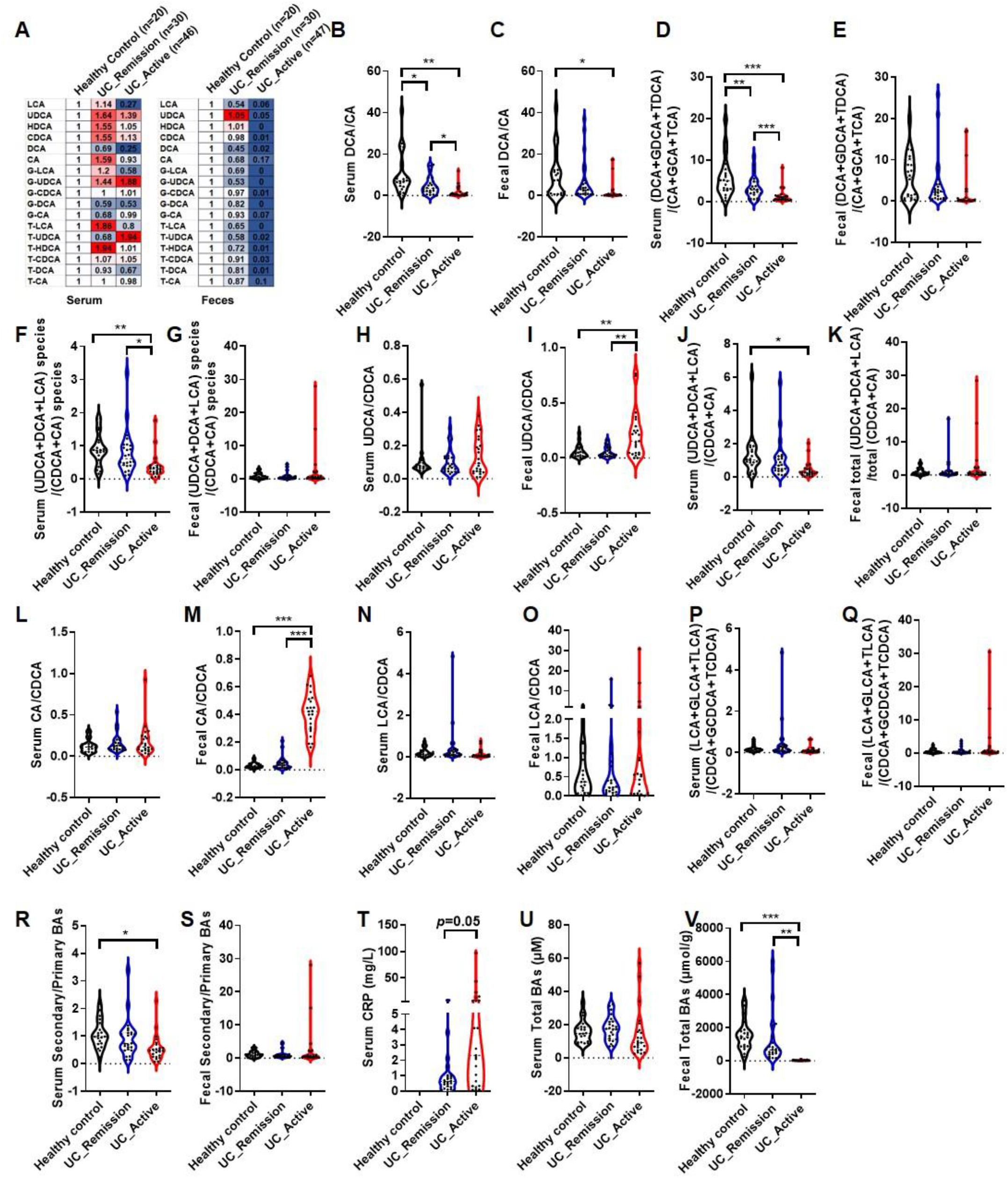
Ulcerative colitis is associated with alternations in bile acids ratios. (A) Normalized average bile acid levels in serum and feces of patients was illustrated by a heatmap. (B-S) Calculated ratios of bile acids in serum and feces. (T) Serum CRP was detected by human CRP assay kit. (U-V) Total bile acid in serum and feces were detected by total bile acid kits. Data are presented as violin plots. ***p < 0.001, **p < 0.01, *p < 0.05; Kruskal-Wallis test.

### Altered bile acid metabolism is associated with altered gut microbiota metabolic function

To ask how exactly microbiota mediated bile acid transformation changed in UC pathology, metagenomic sequencing of fecal samples from healthy control, UC active and UC remission patients was conducted to investigate the relationship the bile acid profiles and intestinal microbiota. Our results showed *Clostrium scindens, Firmicutes bacterium CAG:103, Lachnospiraceae bacterium 5_1_57FAA* and *bacterium LF-3* encoding bile acid-inducible operon (bai) genes involved in secondary bile acid generation. Previous studies showed *Clostrium hiranonis* and *Clostrium hylemonae* also mediate 7α-dihydroxylation^23-25^. Metagenomic sequencing showed UC active patients had lower abundance of *Clostrium scindens, Firmicutes bacterium CAG:103, Lachnospiraceae bacterium 5_1_57FAA, bacterium LF-3, Clostrium hiranonis* and *Clostrium hylemonae* than healthy volunteers (**Figure 3A-F**). In addition, UC remission patients showed higher abundance of *Clostrium scindens, Lachnospiraceae bacterium 5_1_57FAA* and *Clostrium hylemonae* than UC active patients (**Figure 3A-F**). In feces of UC active patients, expression of baiE and baiI was too low to be detected (**Figure 3G-J**), UC active patients showed significantly lower baiCD and baiB expression than healthy volunteers (**Figure 3K and L**), explaining the downregulation of DCA/CA ratio. Moreover, UC active patients showed higher 7α-hsdh expression than healthy volunteer (**Figure 3M**), and proportion of genes encoding enzymes related to bile acid metabolism in microbiota showed UC active patients had larger proportion of genes encoding 7α-hsdh (**Figure 3N**), explaining the upregulation of UDCA/CDCA ratio.

**Figure 3.**
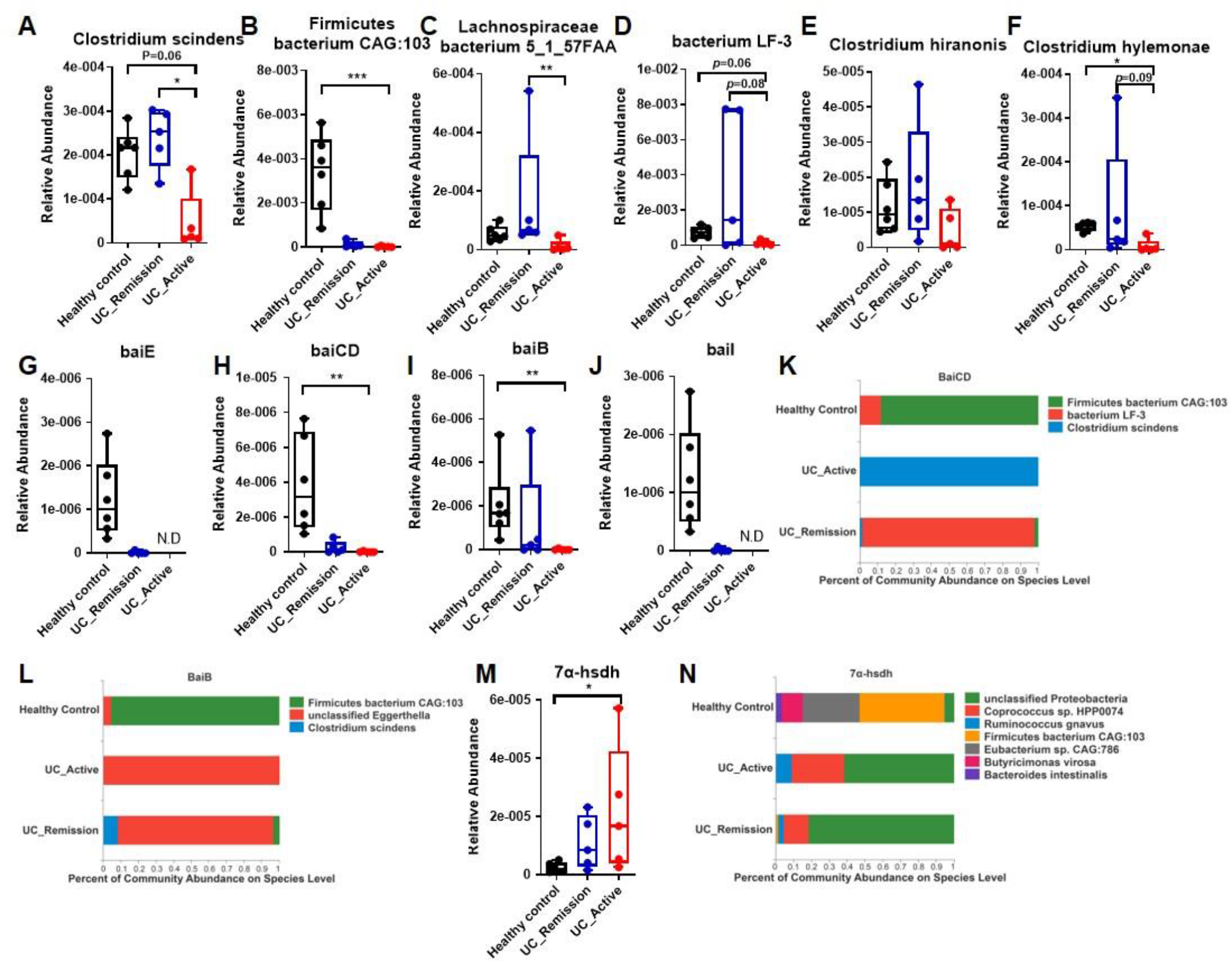
Altered bile acid metabolism is associated with altered gut microbiota metabolic function. (A-F) Relative abundances of bacterial species encoding bacterial enzymes capable of 7α-dihydroxylation in healthy control, UC active and UC remission patients. Relative abundances is calculated by reads number/total reads number of the samples. (G-J) Relative abundances of genes encoding bacterial enzymes capable of 7α-dihydroxylation in healthy control, UC active and UC remission patients, including baiE (I), baiCD (I) and baiB (I). Data are presented as box and whiskers. (K-L) Proportion of genes encoding enzymes related to bile acid metabolism in microbiota of healthy control, UC active and UC remission patients. (M) Relative abundances of 7α-hsdh. (N) Proportion of genes encoding 7α-hsdh in microbiota of healthy control, UC active and UC remission patients. Data are presented as mean. Metagenomics data are presented as box and whiskers. healthy control(n=8), UC active(n=6) and UC remission patients(n=6), ***p < 0.001, **p < 0.01, *p < 0.05; Kruskal-Wallis test. N.D means not detected.

In contribution analysis of baiCD based on annotated species, *Firmicutes bacterium CAG:103* largely accounted for baiCD abundance in healthy volunteers while Clostrium scindens was mainly responsible for total baiCD abundance in UC active and UC remission patients (**Figure 3K**). In addition, various taxa, such as *Lachnospiraceae bacterium 5_1_57FAA, Clostrium scindens* and unclassified *Eggerthella*, contributed to baiB abundance in healthy volunteers and UC remission patients while unclassified *Eggerthella* was the only contributor of genes encoding baiB in UC active patients (**Figure 3L**). Bacteria in *Proteobacteria* and *Enterobacteriaceae* were dominant contributors of genes encoding 7α-hsdh, and in UC active patients and healthy patients, the dominant contributors shared differences (**Figure 3N**). In conclusion, UC active patients showed lower abundance of Bai and taxa encoding Bai. Loss of *Firmicutes bacterium CAG:103* in UC active patients might result in the inhibition of 7α-dihydroxylation in UC active patients. UC active patients showed higher abundance of 7α-hsdh and taxa encoding α-hsdh. This might result from the increase of bacteria in *Proteobacteria*, E.coli, *Ruminococcus gnavus* and *Coprococcus sp. HPP0074* which highly express genes encoding α-hsdh in UC active patients (**Figure S3**).

### Specific bacteria contribute to bile acid metabolic alternation in patients

PCR analysis proved lower abundance of *Clostridium scindens, Firmicutes bacterium CAG:103* and *bacterium LF-3* in UC active patients than in healthy volunteers (**Figure 4A-C**). Abundance of *Clostridium scindens, Firmicutes bacterium CAG:103* and *bacterium LF-3* was higher in UC remission patients than in UC active patients (**Figure 4A-C**). However, no significant positive correlation was observed between the *Clostridium scindens* abundance and serum or fecal (DCA+GCA+TDCA)/(GA+GCA+TCA) ratios (**Figure 4D**). This implied 7α-dihydroxylation inhibition in UC active patients depends on loss of other bacteria except *Clostridium scindens* and *Clostridium scindens* is not dominant bacteria mediating 7α-dihydroxylation in intestine. In addition, *Firmicutes bacterium CAG:103* and bacterium LF-3 abundance was correlated with serum and fecal (DCA+GCA+TDCA)/(GA+GCA+TCA) ratios in UC active patients and UC remission patients (**Figure 4D**). *Lachnospiraceae bacterium 5_1_57FAA* abundance was too low to detect. These results suggested changes of *Firmicutes bacterium CAG:103* and *bacterium LF-3* rather than Clostridium scindens might be the major bacteria influencing (DCA+GCA+TDCA)/(GA+GCA+TCA) ratios alternations in UC patients.

**Figure 4.**
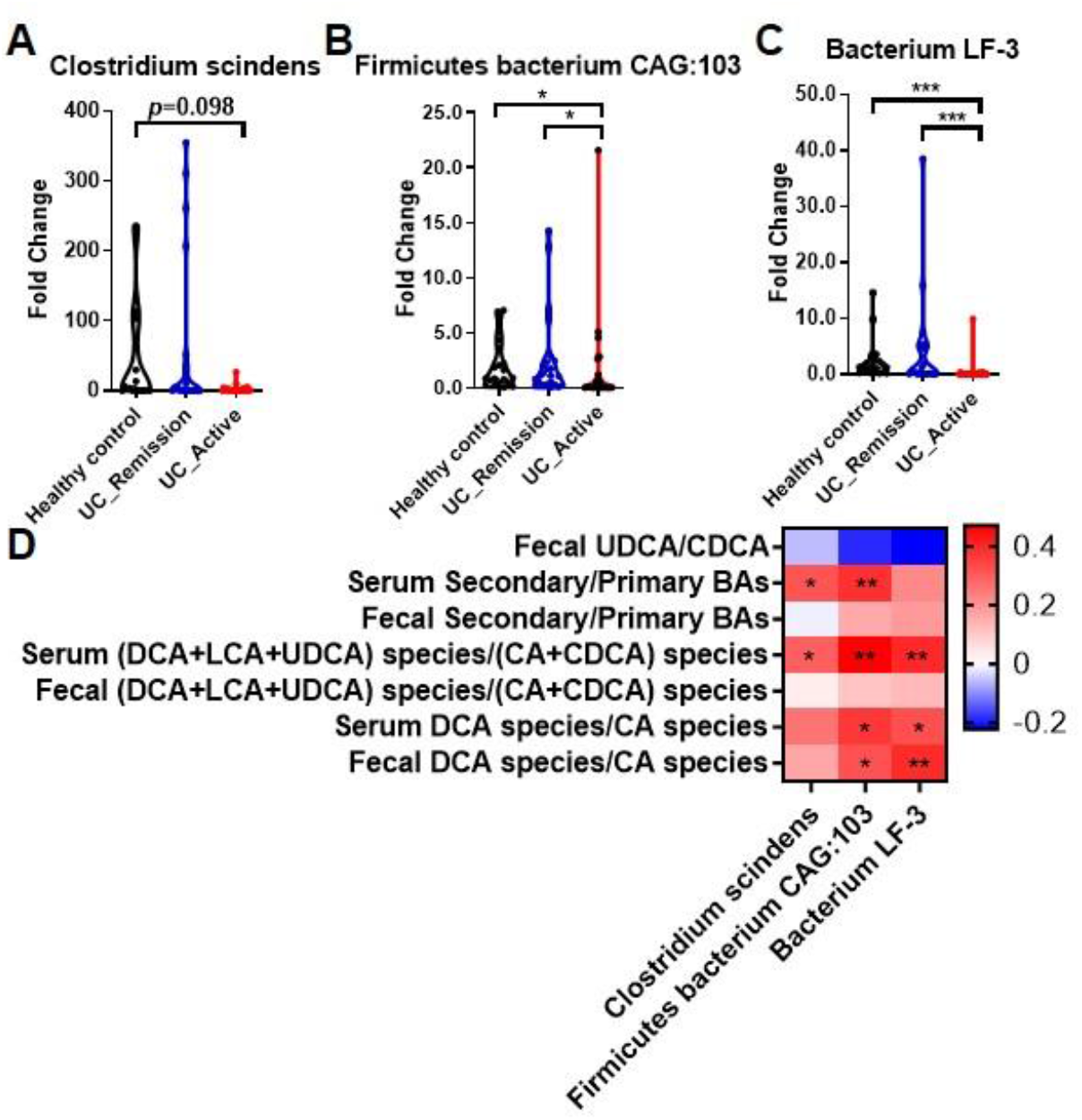
Specific bacteria contribute to bile acid metabolic alternation in patients. (A-C) The fold change of Clostridium scindens, Firmicutes bacterium CAG:103 and Bacterium LF-3 in healthy control, UC active and UC remission patients analyzed by qPCR. (D) Heatmap of Spearman correlation r value between bile acids ratios and the Clostridium scindens, Firmicutes bacterium CAG:103 and Bacterium LF-3 abundances. UC active (n=23) and UC remission patients (n=24). Data are presented as violin plots. Healthy control (n=20), UC active (n=23) and UC remission patients (n=24). ***p < 0.001, **p < 0.01, *p < 0.05; Kruskal-Wallis test.

### Prediction of active UC and remission outcome by calculation of serum and fecal bile acids ratios combined with serum CRP

We further conduct a ROC analysis to test if bile acid composition ratio or related strains were applicable for prediction of UC outcome. In comparable to recognized Ruminococcaceae and Ruminococcus gnavus group which have been proposed to be correlated with UC pathology, DCA producing species including *Clostridium scindens, Firmicutes bacterium CAG:103* and *bacterium LF-3* were more sensitivity in prediction of UC and remission outcome, as evidenced by higher AUC value of ROC curve (**Figure S4, Figure 5A and B**).

**Figure 5.**
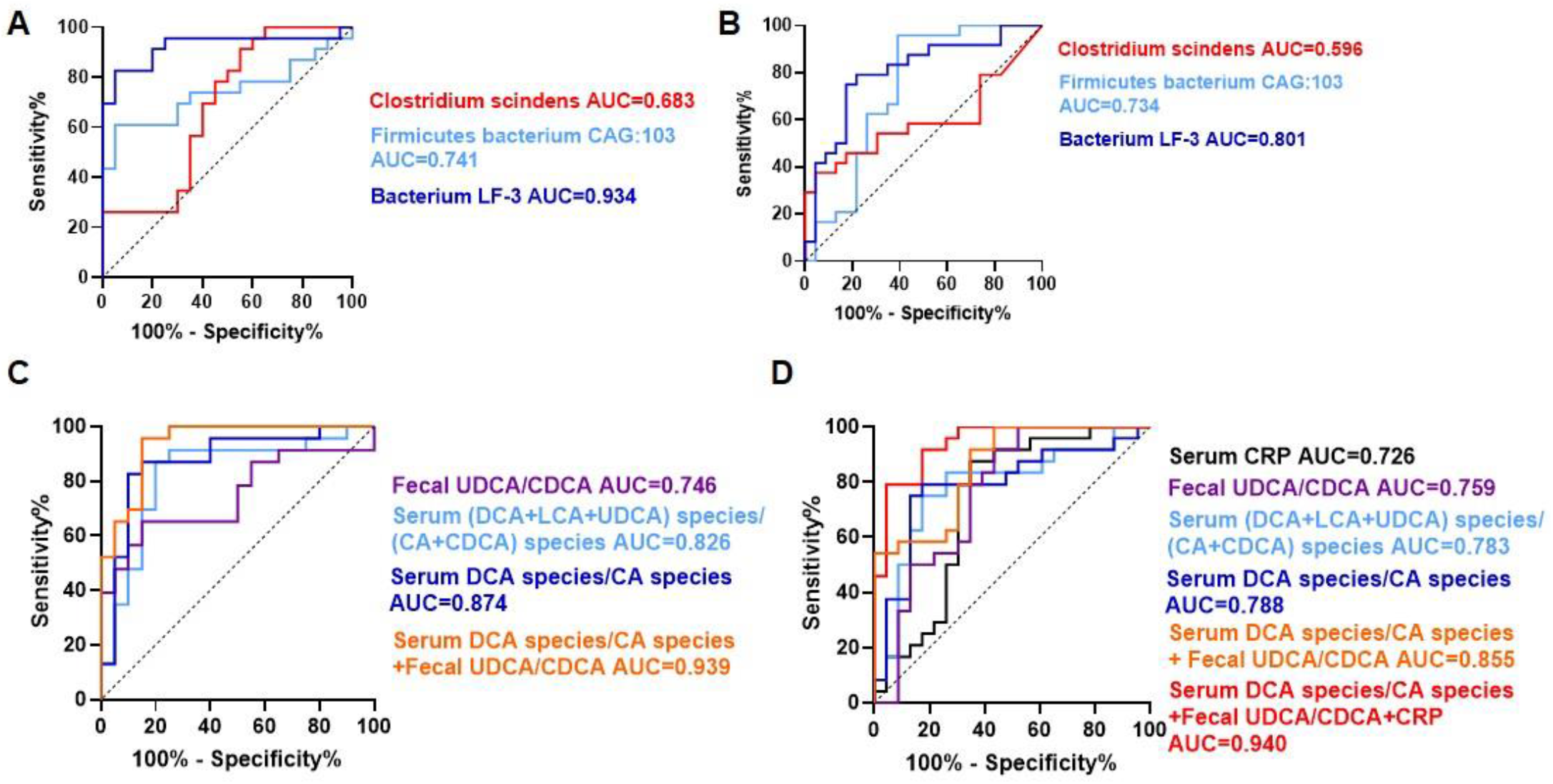
Prediction of active UC and remission outcome by calculation of bacteria abundance and bile acids ratios. (A) ROC curves for UC active patients and healthy controls based on Clostridium scindens, Firmicutes bacterium CAG:103 and Bacterium LF-3. (B) ROC curves for UC remission and UC active patients based on Clostridium scindens, Firmicutes bacterium CAG:103 and Bacterium LF-3. (C) ROC curves for UC active patients and healthy controls based on bile acids ratios. (D)ROC curves for UC remission and UC active patients based on bile acids ratios. Data are presented as violin plots. Healthy control (n=20), UC active (n=23) and UC remission patients (n=24). ***p < 0.001, **p < 0.01, *p < 0.05; Kruskal-Wallis test..

For differentiating UC active patients from normal, bile acid composition ratios were also highly sensitivity in prediction disease states, the prediction sensitivity of fecal UDCA/CDCA ratios levels was 0.746, and totally calculated secondary/primary bile acid species and all DCA forms and CA forms, brought out by (DCA+GDCA+TDCA+LCA+GLCA+TLCA) /(CA+GCA+TCA+CDCA+GCDCA+TCDCA) and (DCA+GDCA+TDCA)/(CA+GCA+TCA) ratios, showed sensitivity values of 0.826 and 0.874, respectively. More specific, the binary logistic regression models concerning the serum (DCA+GDCA+TDCA)/(CA+GCA+TCA) and fecal UDCA/CDCA ratios, showed an sensitivity value of 0.939 (**Figure 5C**).

For differentiating UC remission patients from UC active patients, in comparable to serum CRP, which was invalid differentiated UC remission patients from UC active patients with an sensitivity value of 0.726, the prediction sensitivity of fecal UDCA/CDCA, serum totally calculated secondary/primary bile acid species and all DCA forms and CA forms showed sensitivity values of 0.759, 0.783 and 0.788, respectively. In spite this, the binary logistic regression models concerning the serum serum (DCA+GDCA+TDCA)/(CA+GCA+TCA) ratio, fecal UDCA/CDCA ratio and serum CRP level brought out a prediction sensitivity value of 0.940 (**Figure 5D**).

## DISCUSSION

In this work, we focused on the regular pattern of bile acid metabolism to assess biomarkers distinguishing disease processes in healthy versus UC-remission state and UC-active state groups. In particular, we aimed to quantitative discriminate the UC-remission state and UC-active state groups due to the lack of clinical non-invasive staging diagnostic markers, by investigating the longitudinal change of bile acid profile, bile acid producing gut microbial communities, and bile-acid production strains bias in healthy volunteers and UC patients.

Despite individual differences in absolute bile acid concentrations which may be caused by diet and instrument deviation, composition ratio of different bile acid species remains relative comparatively stable in healthy volunteer group. Moreover, we found composition ratio of DCA species to CA species (DCA/CA), UDCA/CDCA and CA/CDCA were markedly distinctive than serum CRP in healthy versus UC and UC-remission state versus UC-active state groups, we further validated by ROC analysis and found that combined CRP, bile acid and related gut microbiota significantly improved prediction accuracy of active UC and remission outcome.

This work linked bile acid metabolic dysregulation pattern and altered strains with UC pathological activity, yet there are still several vital issues need to be further addressed. For instance, future efforts are imperative for valid confirmation of bile acid as staging biomarkers, strain-specific bile-acid production capabilities of individuals may also be influenced by population diversity of intestinal flora. Nevertheless, we proposed novel insights into the crosslink between metabolome disorder, altered microbiota and disease states, and pinpointed specific bile acid metabolism pathways (ratio and relevant species) that may serve as diagnose and staging biomarkers.

## Data Availability

All data produced in the present study are available upon reasonable request to the authors

## ACKNOWLEDGEMENTS

This work was financially supported by National Natural Science Foundation of China (grants 81720108032, 81930109, 81973559 and 82173886); the National Key Research and Development Program of China (2021YFA1301300); Overseas Expertise Introduction Project for Discipline Innovation (G20582017001) and Sanming Project of Medicine in Shenzhen (grants SZSM201801060 to H. Hao).

## AUTHOR CONTRIBUTIONS

Haiping Hao, Hong Shen and Lijuan Cao designed the study; Lijuan Cao and Yun Wang performed the most experiments and collected and analyzed the data; Lu Zhang collected biosamples, and other authors are all contributive to the experiments; Lijuan Cao, Haiping Hao and Yun Wang wrote and revised the manuscript; Lijuan Cao and Yun Wang contributed equally to this manuscript.

## MATERIALS & CORRESPONDENCE

Further information and requests for materials and correspondence may be directed to the lead contact Haiping Hao.

## SUPPLEMENTARY MATERIALS

**Supplementary Figures S1-S4 and Table S1-S3**.

## FIGURES and FIGURE LEGENDS

**Supplementary Figure S1.**
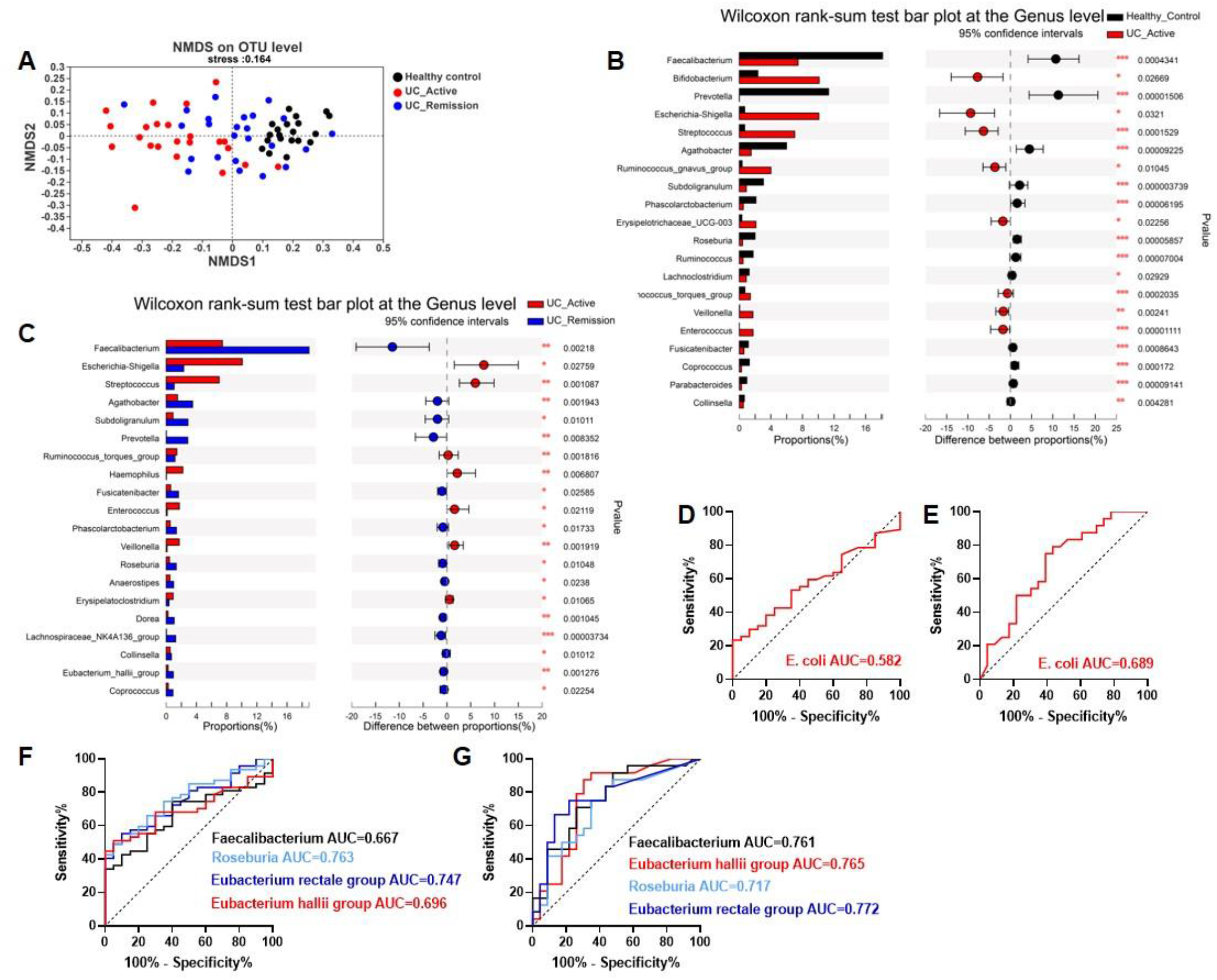
Ulcerative colitis altered the intestinal microbial structure. (A) NMDS of unweighted UniFrac distances of microbiota composition in all groups at OTU levels. (B) The relative abundance of taxa at the genus level in UC active patients and healthy controls. (C) The relative abundance of taxa at the genus level in UC active patients and UC remission patients. (D) Receiver operating characteristic (ROC) curves for UC active patients and healthy controls based on E. coli abundance. (E) ROC curves for UC remission and UC active patients based on E. coli abundance. (F) ROC curves for UC active patients and healthy controls based on Faecalibacterium, Eubacterium rectale group, Roseburia and Eubacterium hallil. (G) ROC curves for UC remission and UC active patients based on Faecalibacterium, Eubacterium rectale group, Roseburia and Eubacterium hallil. Data are presented as violin plots. Healthy control (n=20), UC active (n=23) and UC remission patients (n=24). ***p < 0.001, **p < 0.01, *p < 0.05; Kruskal-Wallis test.

**Supplementary Figure S2.**
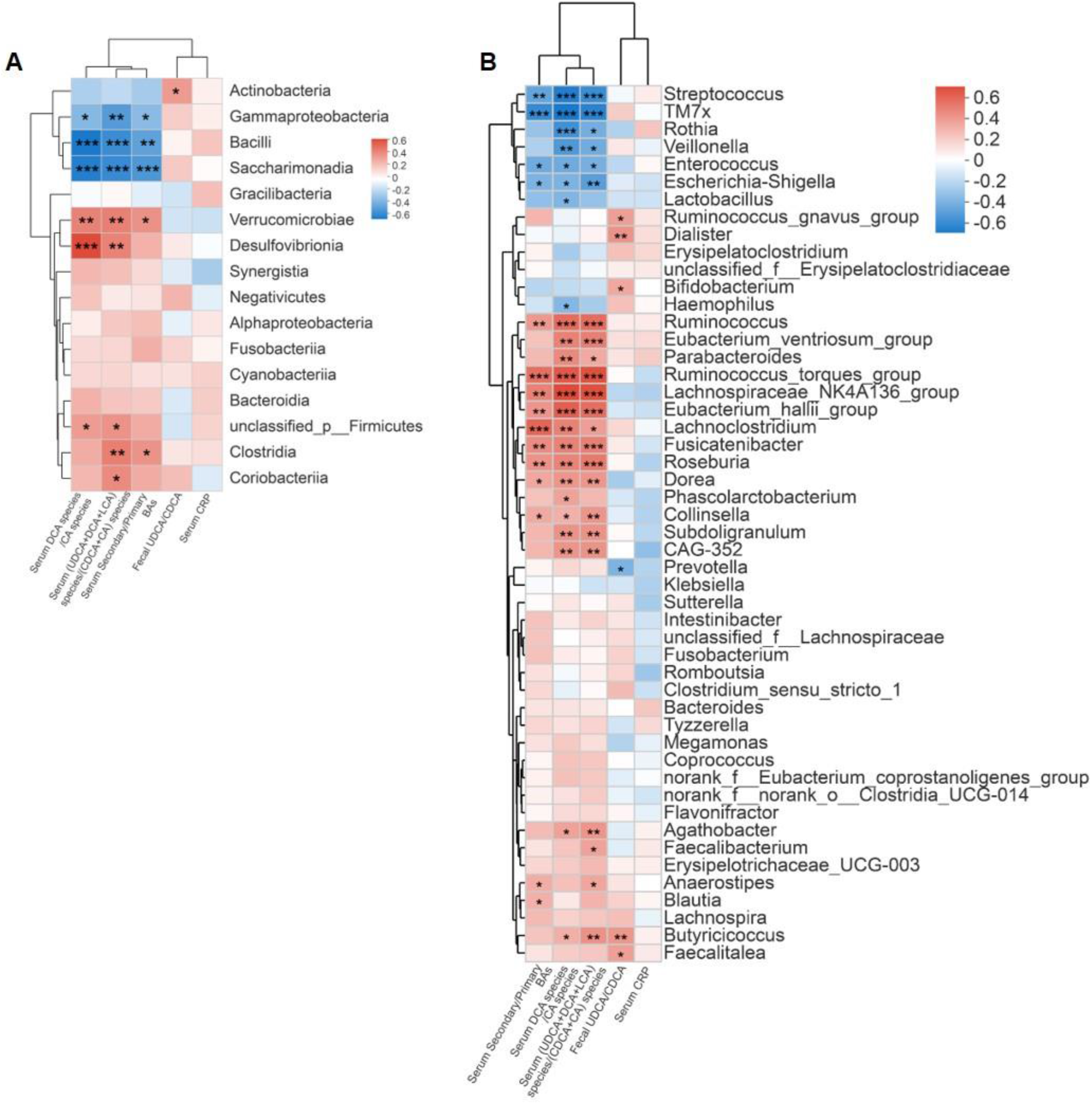
The gut microbial composition is correlated with bile acid ratios. (A-B) Heatmap of Spearman correlation r value between bile acids ratios and taxa at the order and genus level. UC active (n=23) and UC remission patients (n=24). ***p < 0.001, **p < 0.01, *p < 0.05.

**Supplementary Figure S3.**
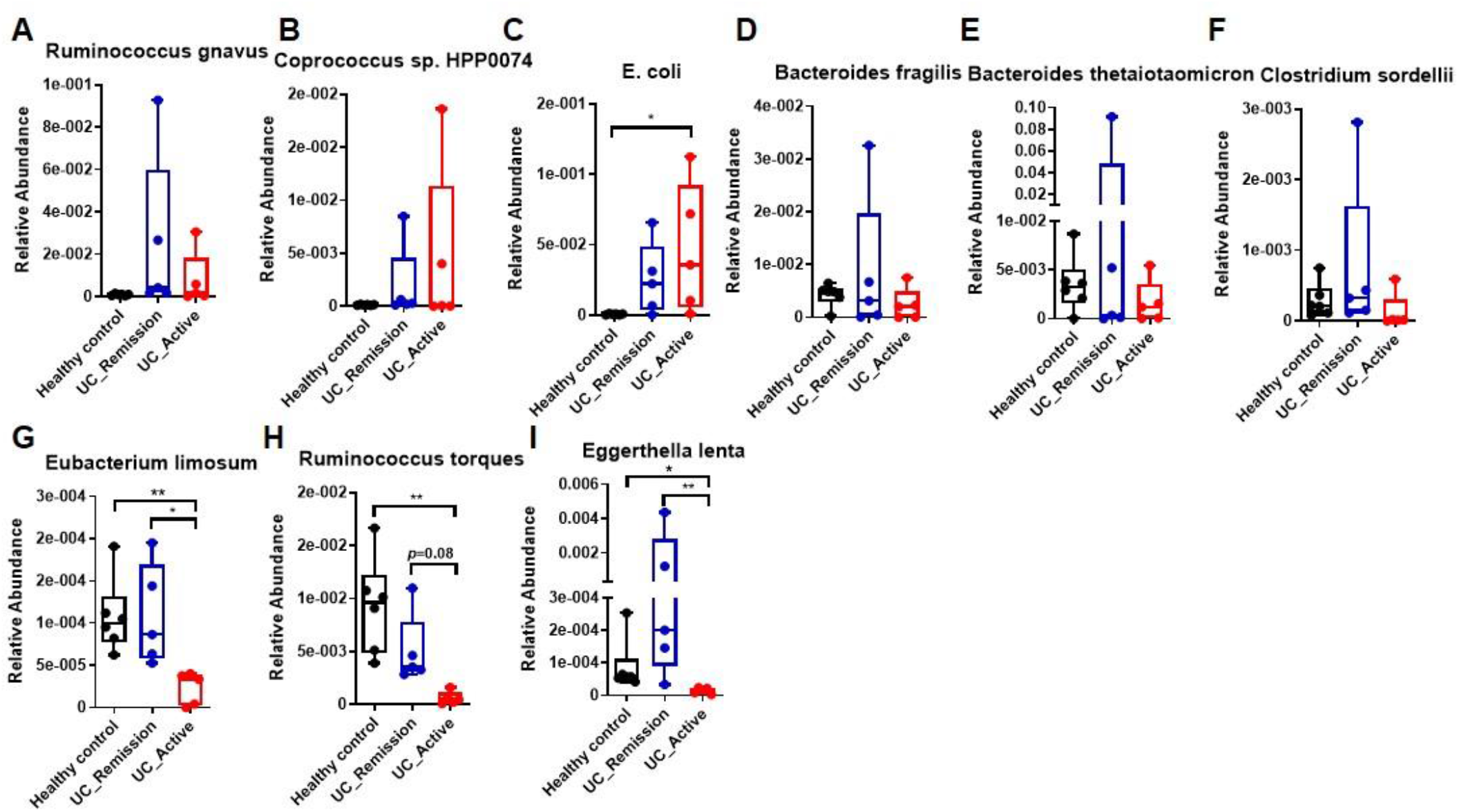
Relative abundances of genes encoding bacterial enzymes of 7α-hsdh capable of 7α-dehydrogenation in healthy control, UC active and UC remission patients. Metagenomics data are presented as box and whiskers. healthy control(n=8), UC active(n=6) and UC remission patients(n=6), ***p < 0.001, **p < 0.01, *p < 0.05; Kruskal-Wallis test. N.D means not detected.

**Supplementary Figure S4.**
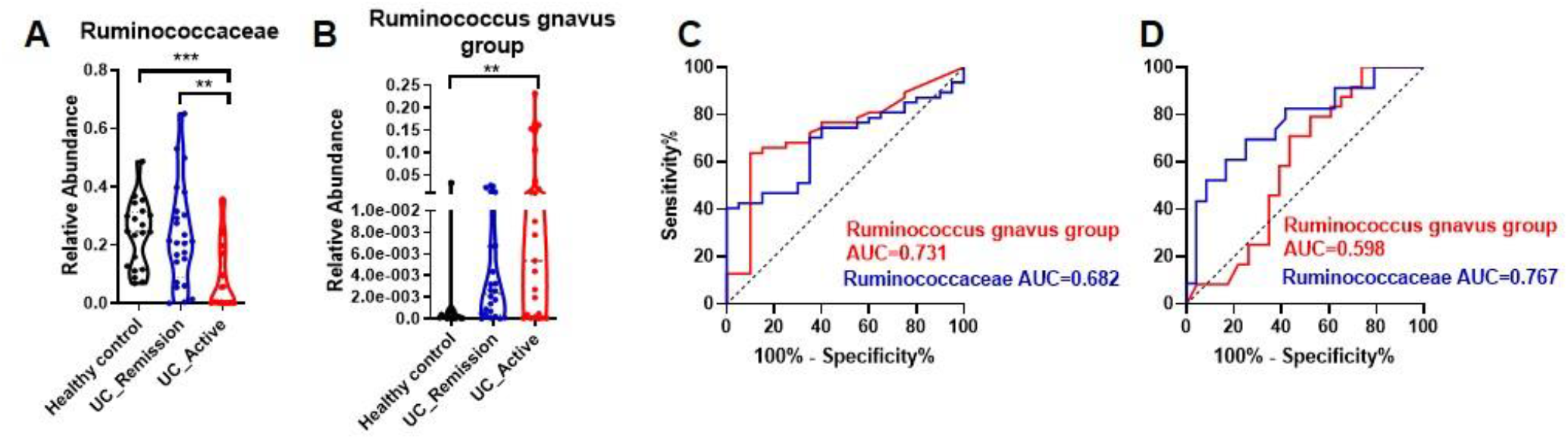
Prediction of active UC and remission outcome by microbiome differences. (A-B) The relative abundance of Ruminococcaceae and Ruminococcus gnavus group in healthy control, UC active and UC remission patients analyzed by 16S rRNA sequencing. (C) ROC curves for UC active patients and healthy controls based on Ruminococcaceae and Ruminococcus gnavus group. (D) ROC curves for UC remission and UC active patients based on Ruminococcaceae and Ruminococcus gnavus group. Data are presented as violin plots. Healthy control (n=20), UC active (n=23) and UC remission patients (n=24). ***p < 0.001, **p < 0.01, *p < 0.05; Kruskal-Wallis test.

**Supplementary Table S1.** Study population.

| Characteristics | Healthy Control (n=20) | UC Remission (n=24) | UC Active (n=23) |
| --- | --- | --- | --- |
| Male/female (%) | 5/15<br>(25/75) | 9/15<br>(37.5/62.5) | 9/14<br>(39.1/60.9) |
| Median (IQR) age (years) | 25 (24.75-26) | 46.5 (37.75-54.25) | 45 (32.5-52) |
| Median (IQR) duration of disease (years) | - |  |  |
| Disease extent, n (%) | - | 23(100%) | 23(100%) |
| E1 proctitis | - | - | - |
| E2 left-sided colitis | - | 17 | 12 |
| E3 pancolitis | - | 7 | 11 |
| Mayo endoscopy subscore, n (%) | - | 24(100%) | 23(100%) |
| Mayo 0 | - | 13(54.1%) | - |
| Mayo 1 | - | 11(45.9%) | - |
| Mayo 2 | - | - | 10(43.5%) |
| Mayo 3 | - | - | 13(56.5%) |
| Concomitant drug treatment, n (%) | 0 (0%) | 5(20.8%) | 23(100%) |
| 5-Aminosalicylic acid | - | 5(20.8%) | 23(100%) |
| Azathioprine/6-mercaptopurine | - | - | - |
| Corticosteroids | - | - | - |
| Immunosuppressants | - | - | - |

**Supplementary Table S2.**
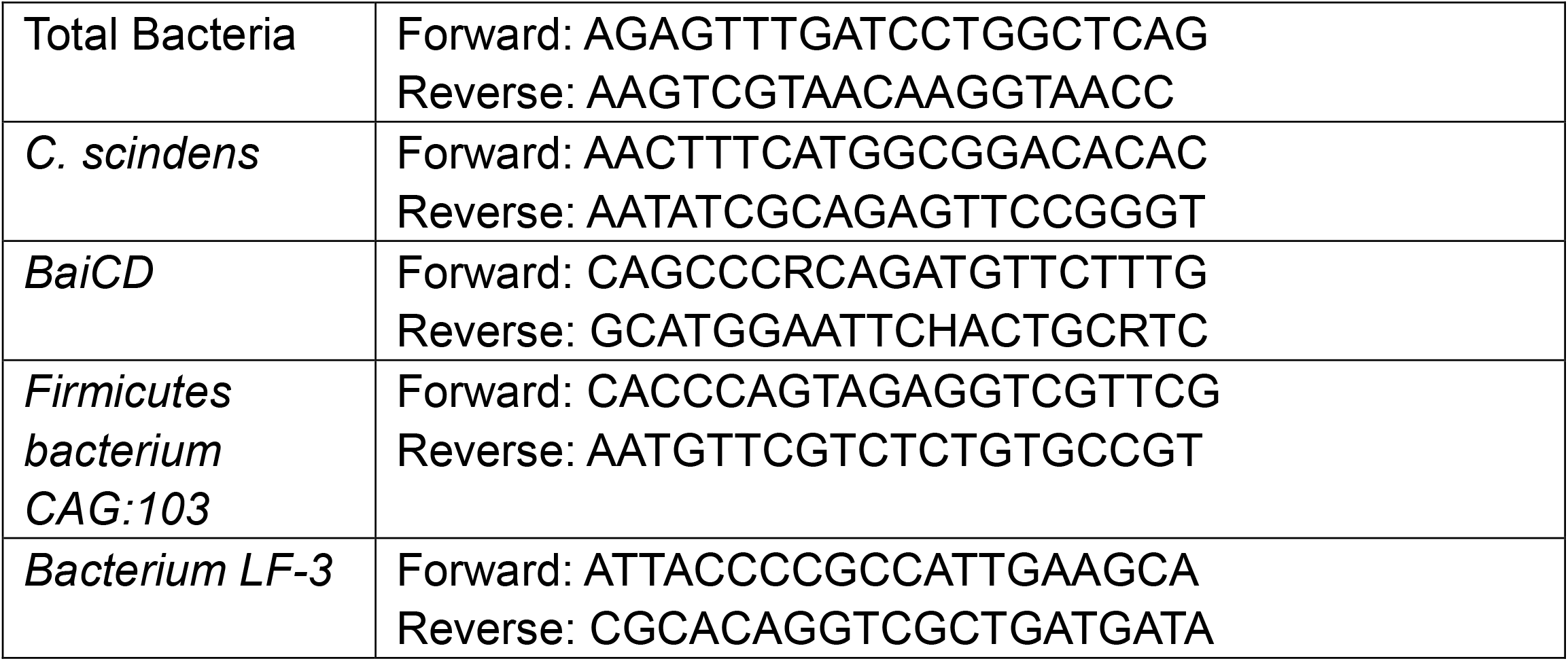
Sequences of primers used in this study.

## METHODS

### CONTACT FOR REAGENT AND RESOURCE SHARING

Further information and requests for reagents may be directed to, and will be fulfilled by the Lead Contact Haiping Hao. All unique reagents generated in this study are available from the Lead Contact without restriction or require a completed Materials Transfer Agreement if there is potential for commercial application.

### CLINICAL STUDY DESIGN AND CHARACTERISTICS

#### Sample collection

Subjects were between 19 and 65 years of age (mean age 39.4±12.9 years) and 60% were female, characteristics of patients see to **Supplementary Table S1**. Stool, urine and blood were collected at an empty stomach in the morning for quantification of biochemical indexes, cytokines, chemokines, bile acids and microbiome profiling.

#### Inclusion and exclusion criteria

Eligibility criteria and staging diagnosis for UC patients were assessed by clinical, endoscopic, and histological diagnosis of ulcerative colitis according to ‘Second European evidence-based Consensus on the diagnosis and management of ulcerative colitis’ (ECCO, 2012). All patients included were chronic relapsing for more than one year, staged into active stage and remission stage by modified Mayo index and Mayo endoscopy scoring. Active-stage UC patients with a course of no less than 4 weeks and free of previous therapy were included, and remission-stage patients were no attack and therapy-naïve for at least 4 weeks.

All patients were excluded from bacterial dysentery, rectitis, severe other diseases (Cardiovascular and cerebrovascular diseases, hepatobiliary diseases, disability or other serious diseases affecting survival), severe complications (local stenosis, intestinal obstruction, intestinal perforation, toxic colonic dilatation, massive hemorrhage, colon cancer or rectal cancer) and history of alcohol and drug abuse. Women patients were excluded from pregnant or lactating.

#### Ethics statement

This study was approved by the ethic committee of Affiliated Hospital of Nanjing University of Traditional Chinese Medicine (2016NL-028-03). Inclusion of healthy volunteers and patients were conducted according to the principles expressed in the Declaration of Helsinki. Written assent was obtained for all patients before any material was taken.

### METHOD DETAILS

#### Quantification of Bile Acids

LC-30A Shimadzu LC system (Shimadzu, Japan) coupled with SIEX Triple Quad™ 5500 system (Sciex, Framingham, MA, USA) was applied, chromatographic separation was performed by a ZOEBAX Eclipse Plus C18 column (2.1×150nm, 3.5μm) (Agilent Technologies, USA). The mobile phase consisted of (A) 1.3 mmol/L ammonium acetate in water (pH adjusted to 6.8 with ammonium hydroxide) and (B) acetonitrile. The flow rate was set at 0.2 mL/min, and the binary gradient elution procedure was performed as follows: initial retentive 20% of solvent B for 5 min, linear gradient 20-25% of solvent B from 5 to 10 min, retentive 25% of solvent B from 10 to 20 min, linear gradient to 55% solvent B from 20 to 25 min and maintained to 29 min, returned to the initial 20% of solvent B in 2 min and held on until 34 min, the injection volume was 5 μL. The mass spectrometer was performed in a full negative scan mode from m/z 50 to 1000 and was operated in SIM mode, reference standards were used to distinguish the isomeric bile acids [M-H]^-^ from each other based on their chromatographic behavior. Analyst 1.4.2 software (Sciex) was used for system control, PeakView software for data exploration and quantification data were acquired with MultiQuant software 2.0.

#### 16S rRNA gene sequencing

Total bacteria genomic DNA in stool samples was extracted by an Omega Biotek Soil DNA Kit (Omega Biotek, Norcross, GA, USA) and the procedures were according to the manufacturer’s instructions. The concentrations and purifications of the bacteria genomic DNA were determined by a NanoDrop 2000 UV-vis spectrophotometer (Thermo Scientific, Wilmington, USA). 1% of agarose gel electrophoresis was used to guarantee the DNA quality. The V3–V4 region of bacterial 16S rRNA gene was amplified with the region-specific primers. The former primer was 338F (5’-ACTCCTACGGGAGGCAGCAG-3’) and the reverse primer was 806R (5’-GGACTACHVGGGTWTCTAAT-3’). TransStart Fastpfu DNA Polymerase (TransGen, Beijing, China) was used. The PCR components consisted of 4 μL of FastPfu Buffer (5×), 2 μL of dNTPs (2.5 mM), 0.8 μL of Forward Primer (5 µM), 0.8 μL of Reverse Primer (5 µM), 0.4 μL of FastPfu Polymerase, 0.2 μL of BSA, 1μL template DNA(10 ng/μL) and 10.8 μL of ddH2O. An ABI GeneAmp 9700 instrument (Applied Biosystems) was used to conduct PCR reaction. The PCR cycling was composed of an initial denaturation at 95 °C for 3 min, followed by 28 cycles of denaturation at 95°C for 30 s, annealing at 55 °C for 30 s and extension at 72 °C for 30 s and finally an extension of 10 min at 72 °C. PCR products were obtained by 2% of agarose gel electrophoresis and were purified with a AxyPrep DNA Gel Extraction Kit (Axygen Biosciences, CA, USA). After ultrasonic fragmentation, library preparation and cluster preparation, paired-end sequencing was conducted on an Illumina MiSeq platform (Illumina, CA, USA) in Shanghai Majorbio Bio-pharm Technology Co., Ltd. Raw reads were demultiplexed, quality-filtered by Trimmomatic. Paired-end reads were assembled with FLASH 1.2.11. All data processing procedures were performed on Majorbio Cloud Platform (http://www.majorbio.com). Sequences were clustered into an operational taxonomic unite (OTU) at 97% similarity with UPARSE 7.0 and chimeric sequences were identified and removed with UCHIME. RDP Classifier 2.11 was used to carry out taxonomical classification based on SILVA database (silva 132/16s bacteria) with a confident threshold of 0.7.

#### Metagenomic sequencing and annotation

Extraction of bacteria genomic DNA was performed as above. The metagenomic sequencing was carried out by Majorbio Bio-Pharm Technology Co., Ltd. After ultrasonic fragmentation, library preparation and cluster preparation, samples were sequenced on HiSeq 4000. Low-quality sequencing reads were removed and human host reads were filtered out. Contigs were generated by Multiple Megahit strategy. Non-redundant gene profiles were set up by mapping quality-controlled reads to the reference gene catalogue. Gene abundance profiles of taxonomies and KEGG orthologous groups (KOs) were calculated as reads per kilobase per million mapped reads (RPKM) based on gene annotation in the NR and KEGG databases. All data processing procedures were carried out on Majorbio Cloud Platform (http://www.majorbio.com).

#### qPCR

Genomic DNA extractions from cecum contents of mice or stool sample of human were performed by using QIAamp Fast DNA Stool Mini Kit (Cat#51604, QIAGEN, Germany). Concentrations of DNA were determined by a Take3 Trio micro-volume plate (Biotek, Winooski, VT, USA) with a microplate reader (Biotek). For clinical stool sample, real-time PCR was conducted based on a SYBR green mixture on a real time PCR cycler (Bio-Rad). Statistical analysis was performed on Graphpad Prism 6.0. Fold change of bacterial abundance and bile acid ratios were taken log calculation for correlation analysis, and correlation was evaluated by Person correlation coefficients. Statistics involved in analysis were valid data excluding those below the detective limitation. For cecum contents of mice, DNA extraction was subjected to PCR-based detection of the 7a-HSDH-encoding baiCD gene. DNA sample was denatured for 2 min at 94ºC, followed by 35 cycles of 94ºC for 20s, 52ºC for 30 s and 69ºC for 90s, and ending with a 10 min extension cycle at 68ºC. PCR products were separated by 1% agarose gel electorphoresis in a Tris–acetate-EDTA buffer system, stained with Gel-red and visualized by UV excitation. Images were captured using iBright CL1000 System (Invitrogen, CA, USA). Specific primers were designed by Primer 3 Plus and blasted based on NCBI genomic database, or obtained from Primer Bank. Sequences of primers used in this study were shown in **Supplementary Table 2**.

### QUANTIFICATION AND STATISTICAL ANALYSIS

I-Sanger planform was used to analyze sequencing data. α-diversity was assessed by Shannon, Chaos, Sobs and Simpson index. To exhibit β-diversity, distances between different groups were measured by principal co-ordinates (PCoA) analysis and non-metric multidimensional scaling (NMDS) based on unweighted unifrac. Variation analysis between pairwise groups on OTUs levels was conducted by Wilcoxon rank-sum test. Binary logistic regression was carried out by IBM SPSS 20. Receiver operating characteristic (ROC) curves for prediction disease activity was performed by GraphPad Prism 8.0 software

### DATA AND SOFTWARE AVAILABILITY

All software used is available online, either freely or from a commercial supplier and is summarized in the Key Resources Table. Data will be accessible upon publication of the manuscript.

